# *In vitro* Characterization of SARS-CoV-2 Protein Translated from the Moderna mRNA-1273 Vaccine

**DOI:** 10.1101/2022.03.01.22271618

**Authors:** Timothy D. Veenstra, Brad Pauley, Elisha Injeti, Rocco J. Rotello

## Abstract

Extensive research around mRNA vaccines and their proposed utility during the current COVID-19 pandemic resulted in many publications concerning the SARS-Cov-2 spike protein and angiotensin converting enzyme-2 receptor-binding domain of the virus, but none describe the characteristics of the full-length protein obtained from the modified/synthetic mRNA that is part of the Moderna and Pfizer-BioNTech vaccines. In this paper, we provide the first data characterizing the actual proteins produced by mouse and human cells in culture that had been incubated up to 30 minutes with the commercial vaccine produced by Moderna (i.e., Spikevax). The mRNA vaccine continues to produce proteins up to 12-14 days after introduction to the cells. The molecular weight of the SARS-CoV-2 encoded protein ranges from 135-200 kilodaltons depending on the extent of glycosylation.

## 1. Introduction

The current coronavirus (COVID-19) pandemic has resulted in several FDA approved emergency use authorizations (EUA) to slow the spread of severe acute respiratory syndrome coronavirus 2 (SARS-CoV-2) [1-3]. Messenger ribonucleic acid (mRNA) vaccines were at the forefront of these EUAs based on their efficacy and how they could be formulated to enter cells after intramuscular injection [4-6]. RNA delivery to cells had been a challenge for decades, however, the development of lipid nanoparticles and the modification with N1-methyl-pseudouridine greatly enhanced the efficacy of both Moderna’s (Spikevax) and Pfizer-BioNTech’s (Comirnaty) mRNA vaccines [7]. In addition, modification of the mRNA with multiple N1-methyl-pseudouridine bases enhanced the stability of the mRNA enabling it to survive the intra and extracellular environments long enough to be translated into a functional, intact protein [8]. One knowledge gap concerning these vaccines was the length of time the mRNA survives *in vivo* and is translated into protein. Another unknown is the size of the actual protein(s) that is translated and what does the actual protein look like after it is made within mammalian cells in culture. In this study, we evaluated the length of time and the size of the protein that the Spikevax vaccine produces *in vitro*. The results show that the vaccine readily enters the cells and produces a variety of protein isoforms within 24 hours. These results suggest the variability in immune response may be due to the translation of different protein isoforms that cause differences in the antibodies produced within the vaccinated or COVID-19 infected subject.

## 2. Materials and Methods

### 2.1 Cell culture system to express SARS-CoV-2 protein(s) from synthetic mRNA-1273 vaccine (Spikevax)

Vaccine uptake was tested in the normal mouse embryonic fibroblast NIH 3T3 adherent cell line (ATCC, Manassas, VA) and human monocytic U937 cells grown in suspension. At confluence, cells were placed in DMEM or RPMI serum-free media (10 mL/100 cm dish). Two hundred microliters of room temperature Spikevax vaccine (Moderna, Cambridge, MA) was added to the cells, which were then incubated for 30 minutes with slow rocking at room temperature. After 30 minutes, 10% (v/v) fetal bovine serum was added, and the cells were placed in an incubator maintained at 37 °C and 4.5% CO_2_. Due to the nature of the lipid, cholesterol, and polyethylene glycol lipid shell that surrounds the mRNA, no other agent was required to assist the entry of the vaccine into cells. This incubation time did not affect cell viability, which remained above 90% based on trypan blue exclusion and cell counting using a CytoSMART cell counter (CytoSMART Technologies LLC, Skillman, NJ). At selected time points, cells were collected and lysed in Triton lysis buffer containing a protease inhibitor cocktail. The cell supernatants were also collected and stored at -20 °C for further analysis. Protein concentrations were measured using a Bradford microplate assay (Biorad, Hercules, CA).

### 2.2 Enzyme-linked Immunoassay

Enzyme-linked immunoassay (ELISA) reactivity was evaluated against the commercially available SARS-CoV-2 protein (R&D Systems, Minneapolis, MN). Briefly, Immulon™ 2 high protein binding ELISA plates (ICT; Davis, CA) were coated with SARS-CoV-2 proteins at 100 ng/well in phosphate buffered saline (PBS) at 4 °C overnight. Wells were rinsed twice with PBS and blocked using 3% blotting-grade protein blocker (Biorad). Antibodies were incubated in concentration ranges from 1-10 μg/ml and detected using donkey anti-human secondary antibodies, (Jackson ImmunoResearch Laboratories; West Grove, PA) labeled with horse radish peroxidase (HRP) at a 1:5,000 dilution. These secondary antibodies are supplied pre-adsorbed to numerous species’ IgGs to reduce non-specific binding. After incubation with 3,3’,5,5’-tetramethylbenzidine (TMB) substrate for 10 minutes, 50 μl of 2 N sulfuric acid was added and the absorbance of the TMB substrate at 450 nm was measured using a Promega plate reader. The binding of the collected human saliva and REGEN-COV2 antibodies to cell lysates and supernatants was evaluated using an in-house developed ELISA. To test for the presence of autoantibodies, the ELISA was performed by coating the wells of the plate with the mRNA vaccine and probing them with the patient samples (Groups A-D) along with the commercial antibodies. No positive signals were observed in the ELISA showing that vaccinated patients do not develop autoantibodies against the vaccine.

### 2.3 Western blot of proteins from cells incubated with mRNA-1273 (Spikevax)

Cell lysates were quantitated and combined with 4x Laemmli buffer and heated at 100 °C for 5 minutes. The cell lysate (20 μg) was separated using a 6-18% SDS-PAGE gel. Proteins were transferred onto a nitrocellulose membrane under ice for 1 hr at 100 volts. Membranes were blocked using 3% blotting-grade blocker (BioRad) and washed with PBS. To verify the actual molecular weights of the proteins, a mouse monoclonal antibody from R&D Systems, specific to the S2 subunit protein, (met697-Pro1213; cat# 1034617), was used at a concentration of 1 μg/ml diluted in Blotto (Santa Cruz Biotechnology, Dallas, TX). In addition, western blotting with clarified saliva antibodies, was performed with samples diluted 1:3 in blotting-grade blocker, while the REGEN-COV humanized IgG_1_ antibodies were dissolved at 10 μg/ml in blotting-grade blocker. Secondary antibodies were used at a dilution of 1:2500 in 3% blotting-grade blocker. Enhanced chemiluminescence (ECL) was employed to detect proteins, using an Azure 600 imager (Azure Biosystems, Dublin, CA).

### 2.4 Human saliva samples

Saliva samples were obtained from individuals, aged 50-65 years of age, who were (Group A) unvaccinated and contracted COVID-19; (Group B) vaccinated, had not contracted COVID-19, and tested negative for the virus; (Group C) unvaccinated, had not contracted COVID-19, and had no disease-related symptoms between September 2020 and October 2021; (Group D) vaccinated and had contracted COVID-19 based on a rapid antigen test. All samples were acquired with approval from the Cedarville University Institutional Review Board.

Saliva samples were tested from donors and volunteers at random time points using an ELISA. Samples were collected in a collection vial containing 0.05% thimerosal. The samples were immediately centrifuged at 21,000 rpm at 4 °C to remove any particulates and stored at 4 °C. Samples being stored longer than 2 weeks contained a protease inhibitor cocktail (4-(2-aminoethyl)benzenesulfonyl fluoride hydrochloride, aprotinin, bestatin, E64, leupeptin, and pepstatin A; Cell Signaling Technology, Danvers, MA), 10% glycerol and were stored in aliquots at -20 °C.

## 3. Results and discussion

### 3.1 Anti-SARS-CoV-2 monoclonal antibody, human saliva, and REGEN-COV antibody binding to proteins extracted from Spikevax-treated fibroblasts

Twelve saliva samples (3 from each group A-D) were analyzed in duplicate using an ELISA plate coated with the SARS-CoV-2 spike protein. Anti-SARS-CoV-2 monoclonal antibodies (REGEN COV and R&D Systems) were used as positive controls in both the ELISA and western blots (see section 3.2). Saliva obtained from subjects in group A who presented with similar clinical profiles (i.e., unvaccinated, serious illness followed by recovery) had the strongest reactivity to the SARS-CoV-2 protein. Samples taken from Group B (vaccinated and non-COVID-19 infected) and Group D (vaccinated and previously infected with COVID-19) subjects exhibited a similar response in the ELISA (Figure 1), however, it was less than that observed in samples from Group A subjects. Saliva samples from Group C subjects (unvaccinated and non-COVID-19 infected) showed essentially no response, confirming the lack of anti-SARS-CoV-2 spike protein antibodies in these subjects that had not been vaccinated nor infected by COVID-19. Antibodies were purified from Group A saliva samples and isotyped as strong IgG1 and IgG2a using lateral flow strips (Thermo Fisher Scientific, Waltham, MA). While the commercial monoclonal antibodies from R&D Systems and Regeneron (REGEN-COV2) showed positive reactivity in the ELISA, their response was lower than that observed using saliva samples from Group A subjects.

**Figure 1.**
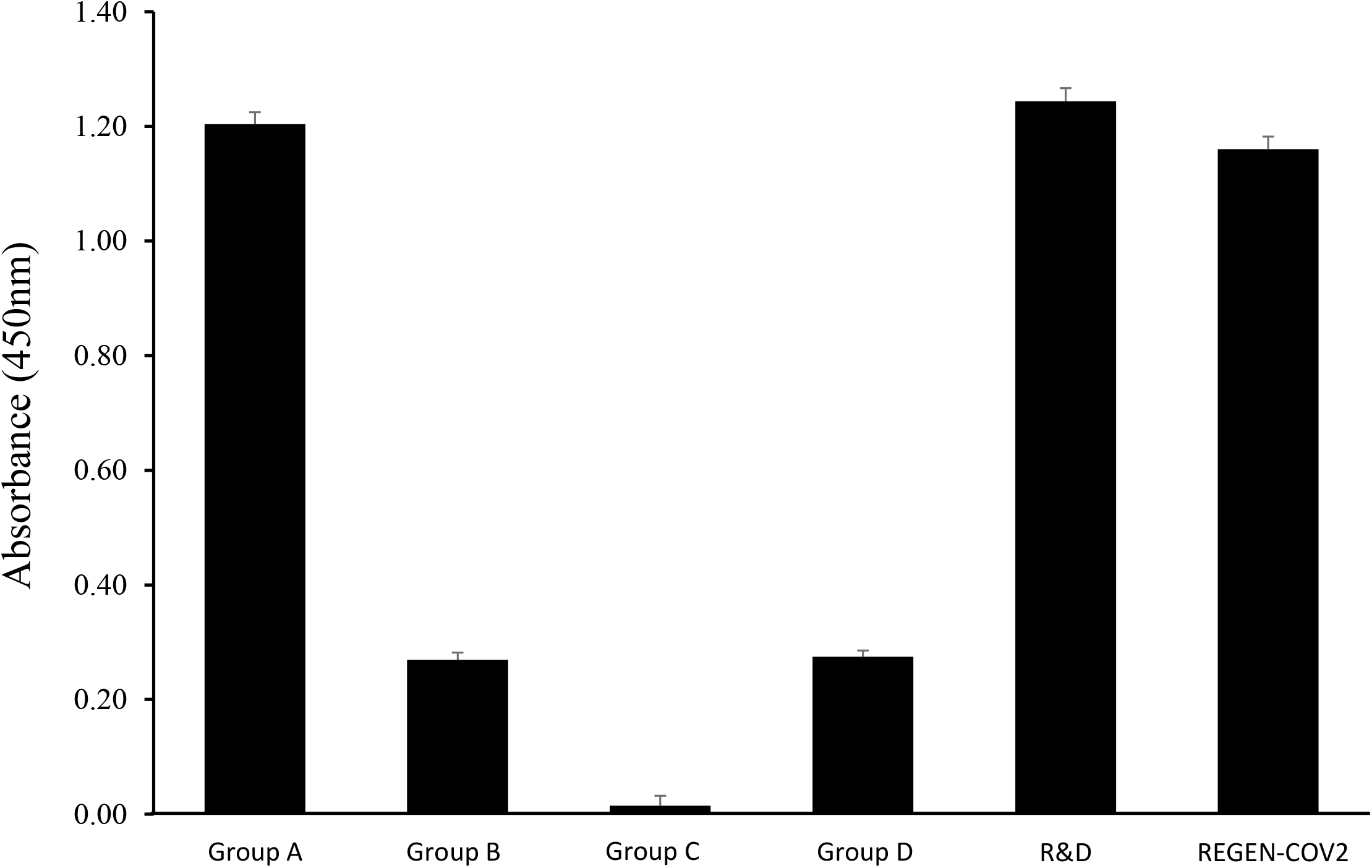
Enzyme-linked immunoassay results showing the binding of antibodies within saliva samples from an unvaccinated, COVID-infected (Group A), vaccinated, non-infected (Group B), unvaccinated, non-infected (Group C), and vaccinated, infected (Group D) individuals. Commercially available antibodies (REGEN COV and R&D Mab) were used as positive controls to the SARS-CoV-2 spike protein.

### 3.2 Western blots of extracts from Spikevax-treated cells

Cell lysates and supernatants were analyzed using western blotting to determine the size of protein expressed from the Spikevax vaccine mRNA (which has not been previously reported in the literature). Cell lysates and supernatants collected at 1, 3, 6, 12 and 24 hrs, and 5, 9, and 15 days, were analyzed using an ELISA and western blotting for presence of Spikevax-synthesized protein. Western blotting using a mouse monoclonal antibody from R&D Systems revealed three prominent bands at a molecular weight of approximately 180 kD (Figure 2), which can be seen most prominently in the cell lysates at 24 hours post-infection with the vaccine. The three bands with distinct molecular weights may arise from differential post-translational modifications (most likely glycosylation) that occurs as the proteins expressed from the mRNA vaccine are processed. SARS-CoV-2 protein expression was detectable in cell lysates within 6 hours of treating the cells with the vaccine (Figure 2). Protein levels peaked at 24 hours and remained detectable over 5 days. No SARS-CoV-2 spike protein was detectable in the NIH 3T3 cell lysates after 12 days. The cell supernatants did not contain any detectable vaccine-induced protein.

**Figure 2.**
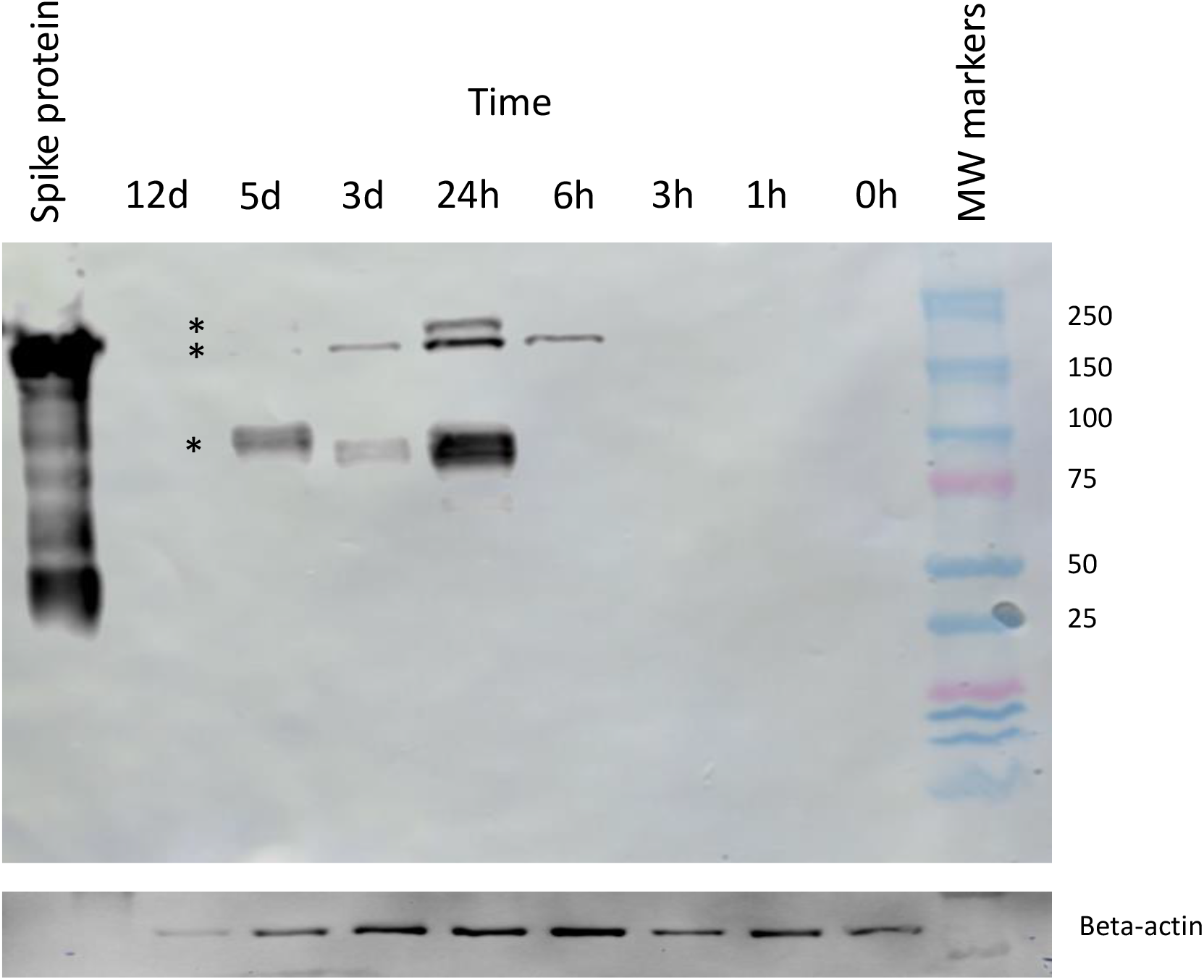
Western blot of NIH 3T3 cell extracts harvested at various time points after Spikevax-treatment, with mouse monoclonal antibody (R&D Systems, Cat^#^ 1034617). Asterisks indicate presence of mature protein and protein being actively post-translationally modified. Mouse monoclonal antibody was used at 1ug/ml blotto to bind Beta actin (R&D systems, Cat # 3700, clone 8H10D10).

The expression of Spikevax was also analyzed in U937 cells (Figure 3). While vaccine protein expression is detectable in lysates obtained from both cell lines, spike protein levels were much lower in the U937 cells compared to NIH 3T3 cells. Detectable protein was not observed in U937 cell lysates until 24 hours after incubation with the vaccine. The numerous isoforms observed in NIH 3T3 cells were also not clearly detected in U937 cells. This reduced expression may be due to U937 cells being a leukemic cancer cell line while NIH 3T3 cells are non-tumorigenic and non-genetically modified [9].

**Figure 3.**
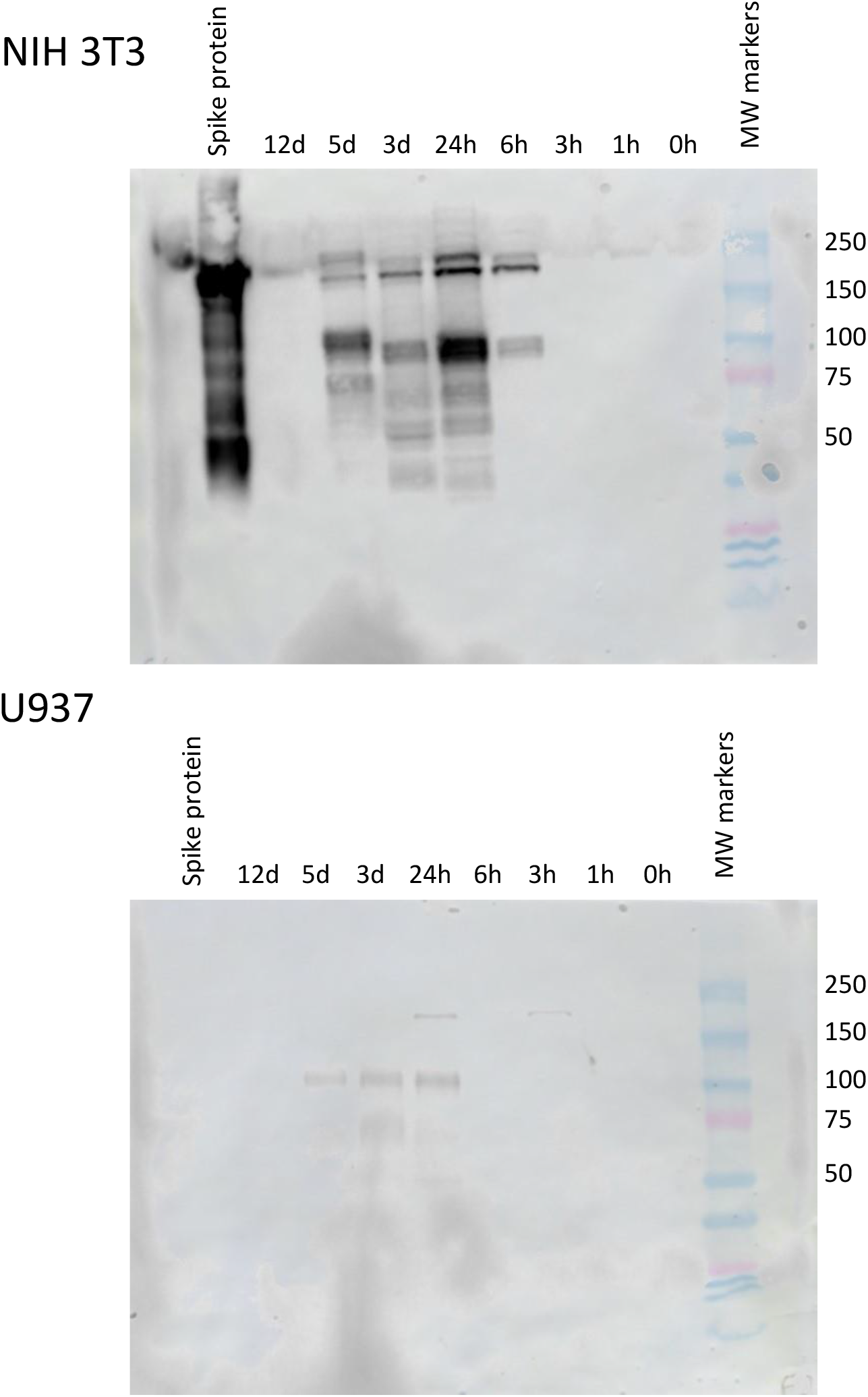
Western blot of NIH 3T3 (top) and U937 (bottom) cell extracts using mouse monoclonal antibody directed against SARS-CoV-2 protein at various time points after treating the cells with Spikevax. The REGEN COV antibody does not bind to the SARS-Cov-2 protein in a western blot format, only in a soluble format, see ELISA data, Figure 1.

## 4. Discussion

Isolating mRNA is technically challenging owing to its susceptibility to degradation by many RNA-degrading enzymes and buffers [10]. This fact makes the ability to deliver an mRNA vaccine in a lipid nanoparticle vesicle through a needle into different cell types a remarkable success. The mRNA vaccine is expressed and post-translationally modified using the machinery akin to any wild-type mRNA that may be normally present within the cell. This study shows that mouse NIH 3T3 cells take up the vaccine and start expressing protein within 6 hours. The protein encoded by the mRNA vaccine is detectable within cell extracts isolated 5 days post-treatment, however, no protein is detectable in the cell supernatants at this time point (ELISA data not shown). In communications with Moderna and Pfizer-BioNTech regarding the proteins expressed by their synthetic mRNA vaccines, each company’s medical information group disclosed that that they had not examined the protein dynamics more than 48 hours post-transfection in cell culture. Owing to its proprietary status, they would not disclose any information related to the nature of the protein that was expressed.

Both Moderna and Pfizer-BioNTech’s mRNA vaccines encode a protein with a predicted molecular weight of approximately 141 kD. The protein contains 22 predicted N-glycosylation and several O-glycosylation sites, making it heavily glycosylated [11]. Full glycosylation of these sites would probably increase the protein’s molecular weight well above 200 kD. In this study, Western blotting using a commercially available mouse monoclonal anti-SARS-CoV-2 spike protein antibody and antibodies from saliva of non-vaccinated individuals, positive for COVID-19 showed three protein bands with molecular weights around 180 kD. While the exact molecular composition of these expression products is not known, the increase in apparent size compared to the predicted size, suggests that the protein expressed from the vaccine is glycosylated. The results show that the protein expressed from the synthetic mRNA vaccine is present in multiple isoforms within the cell. These levels of these isoforms may play a role in the differential response that individuals have to the vaccine.

## Data Availability

All data produced in the present study are available upon reasonable request to the authors.

## Acknowledgments

The author(s) would like to thank Cedarville University for financial support during the research, authorship, and publication of this article.

